# Cardiorespiratory Fitness in Adolescence and Risk of Type 2 Diabetes in Late Adulthood: A Nationwide Sibling-Controlled Cohort Study

**DOI:** 10.1101/2024.11.26.24318038

**Authors:** Marcel Ballin, Viktor H. Ahlqvist, Daniel Berglind, Mattias Brunström, Herraiz-Adillo Angel, Pontus Henriksson, Martin Neovius, Francisco B. Ortega, Anna Nordström, Peter Nordström

**Author notes:** Correspondence: Marcel Ballin. Department of Public Health and Caring Sciences, Clinical Geriatrics, Uppsala University, Husargatan 3 BMC, SE 75122, Uppsala, Sweden.

## Abstract

**Background:** The importance of adolescent cardiorespiratory fitness for long-term risk of type 2 diabetes (T2D) remains poorly investigated, and whether the association is influenced by unobserved familial confounding is unknown.

**Methods:** We conducted a sibling-controlled cohort study based on all Swedish men who participated in mandatory military conscription examinations from 1972 to 1995 around the age of 18, and who completed standardized cardiorespiratory fitness testing. The outcome was T2D, defined as a composite endpoint of diagnosis in inpatient or specialist outpatient care, or dispensation of antidiabetic medication, until 31 December 2023.

**Findings:** 1 124 049 men, of which 477 453 were full siblings, with a mean age of 18.3 years at baseline were included. During follow-up, 115 958 men (48 089 full siblings) experienced a first T2D event at a median age of 53.4 years. Compared to the first decile of fitness, higher fitness levels were associated with a progressively lower risk of T2D. In cohort analysis, the hazard ratio (HR) in the second decile was 0.83 (95% CI, 0.81 to 0.85), with a difference in the standardized cumulative incidence at age 65 of 4.3 (3.8 to 4.8) percentage points, dropping to a HR of 0.38 (0.36 to 0.39; incidence difference 17.8 [17.3 to 18.3] percentage points) in the tenth decile. When comparing full siblings, and thus controlling for all unobserved behavioral, environmental, and genetic confounders that they share, the association replicated, although with attenuation in magnitude. The HR in the second decile was 0.89 (0.85 to 0.94; incidence difference 2.3 [1.3 to 3.3] percentage points), and in the tenth decile it was 0.53 (0.50 to 0.57; incidence difference 10.9 [9.7 to 12.1] percentage points). Hypothetically shifting everyone in the first decile of fitness to the second decile was estimated to prevent 7.2% (6.4 to 8.0) of cases at age 65 in cohort vs. 4.6% (2.6 to 6.5) in full-sibling analysis. The association was similar in those with overweight as in those without.

**Interpretation:** Higher levels of adolescent cardiorespiratory fitness are associated with lower risk of T2D in late adulthood, with clinically relevant associations starting already from very low levels of fitness, and similarly in those with overweight compared to those without. The association replicates, but becomes weaker, after adjusting for unobserved familial confounders shared between full siblings. This suggests that adolescent cardiorespiratory fitness is a robust marker of long-term T2D risk, but that conventional observational analysis may yield biased estimates.

**Funding:** None.

**Research in context:** *Evidence before this study:* Type 2 diabetes is a growing public health issue, affecting at least half a billion people globally. Modifiable factors such as physical activity and the closely related trait cardiorespiratory fitness, which are inversely associated with type 2 diabetes, are declining, particularly among youth. Since these traits track from youth into adulthood, early-life interventions might have important implications for prevention. However, previous studies have typically focused on middle-aged individuals, leaving gaps in understanding whether fitness in youth is associated with type 2 diabetes in the long-term. Moreover, they have not been designed to adequately account for unobserved confounders. Triangulating the evidence across different methods, such as using sibling comparison analysis, would be important to obtain more accurate and reliable estimates of the causal relationship.

*Added value of this study:* In this nationwide sibling-controlled cohort study encompassing more than 1 million young men, of which half a million were full siblings, higher levels of adolescent cardiorespiratory fitness were associated with a substantially lower risk of developing type 2 diabetes up to five decades later. The association was clinically relevant already from low levels of fitness, and it appeared similar in those with overweight as in those without overweight. While the association replicated after adjusting for unobserved familial confounders shared between full siblings, the magnitude of association attenuated by an amount that appeared clinically relevant. For example, the incidence differences between deciles of fitness were about 40% smaller in sibling-comparison analyses as compared to cohort analysis, and the preventable share of type 2 diabetes associated with hypothetical interventions shifting the population-level of fitness was reduced by about one-third.

*Implications of all the available evidence:* Adolescent cardiorespiratory fitness is a strong marker of long-term risk of type 2 diabetes, both in those with and without overweight. These findings render support to large-scale surveillance of fitness from a prevention perspective, and if the findings are confirmed using other lines of causal analysis, they may render support to interventions targeting fitness already from a young age. Yet, these findings also highlight the importance of triangulation for obtaining more reliable evidence of the magnitude of association, and shed light on the pitfalls of conventional observational analysis which may yield biased estimates.

## Introduction

Type 2 diabetes (T2D) is a major public health concern, affecting at least half a billion people in 2019.^1^ In conjunction with the rising prevalence of overweight and obesity which is a key contributor to T2D burden,^2,3^ other modifiable risk factors including physical activity and its closely related trait cardiorespiratory fitness, are trending downwards,^4,5^ particularly in younger age groups^6,7^ and worsening after the recent COVID-19 pandemic.^8–10^ Because these traits track from a young age into adulthood,^11,12^ adolescence may represent an underutilized age group for preventive efforts.

Unfortunately, the current observational evidence linking cardiorespiratory fitness with T2D is primarily based on studies in middle-aged individuals,^13^ and much less is known about the role of fitness in adolescence, including whether it differs between overweight and normal weight individuals.^14^ An important limitation of previous studies is also that they are largely susceptible to confounding, particularly from unobserved familial factors, limiting their possibility to infer causality. Because it is notoriously challenging to address unobserved confounding in conventional observational analysis, triangulating the evidence across different methods would be crucial to ensure reliable evidence that can support public health policy and interventions.^15^

One such approach is family-based analysis, whereby all shared factors, including unobserved behavioral, environmental, and genetic confounders, are inherently controlled by comparing family members (such as siblings) who are discordant for the exposure and outcome.^16^ However, to our knowledge, there has been no previous study employing family-based analysis, such as sibling comparisons, when examining the role of cardiorespiratory fitness in T2D.

Therefore, we performed a nationwide cohort study encompassing over 1.1 million young men tested for cardiorespiratory fitness and who were prospectively followed for up to five decades for T2D incidence. To gauge the validity of the observational findings, we performed sibling-comparison analyses in half a million full siblings, thereby partly addressing the currently unknown influence of unobserved confounding to the association.

## Methods

### Design

This was a registry-based cohort study conducted by cross-linking data from nationwide data sources in Sweden, using the Personal Identification Number, which is unique for all Swedish residents. The study was approved by the Regional Ethical Review Board in Umeå and the Swedish Ethical Review Authority (no. 2010-113-31M), and the need for informed consent was waived. The study is according to the STROBE guidelines.^17^

### Data sources

The study population was obtained from the Swedish Military Service Conscription Register,^18^ and included all men who participated in military conscription examinations from 1972 to 1995 and who had completed cardiorespiratory fitness testing. During this period, conscription around the age of 18 years was mandatory for all Swedish men with few exemptions (i.e., 90% of the male population was conscripted).^18^ Exemptions included specific functional disabilities, routine care needs, specific religious beliefs, or conscripting elsewhere. Using the Multi-Generation Register, we identified all conscripts who were full siblings.^19^ Using the National Patient Register, we collected data on T2D diagnoses from inpatient and specialist outpatient care.^20^ This register was launched in 1964, and from 1987 it has complete coverage of inpatient diagnoses, and since 2001 it also includes data on specialist outpatient care. We collected data on the dispensation of antidiabetic medications using the Prescribed Drug Register, which was launched nationwide in July 2005 and includes data on all drugs dispensed at pharmacies in Sweden,^21^ in an attempt to also capture primary care treatment/diagnosis of T2D which is not covered by the patient register. Deaths were identified using the National Cause of Death Register, which has complete coverage since 1951,^22^ and emigration and socioeconomic data from databases managed by Statistics Sweden (reporting mandated by law).^19,23^

### Derivation of study population

Between 1972 and 1995, a total of 1 249 131 Swedish men were conscripted. From these, we excluded 33 645 (2.7%) with missing cardiorespiratory fitness data, 72 042 (5.8%) with missing data on any covariates, and 19 395 (1.5%) with extreme values recorded for cardiorespiratory fitness (see below). As such, a total of 1 124 049 men with complete data were included in the cohort analysis (90.0% retained) (Supplemental Figure 1). Among these, 477 453 were identified as full brothers (219 304 families) and were subsequently included in the full-sibling analysis.

### Assessment of cardiorespiratory fitness

The exposure was cardiorespiratory fitness, assessed during conscription using a maximal ergometer bicycle test and according to a standardized protocol.^18^ The test results were recorded as watt maximum (W_max_). Briefly, conscripts performed the test after presenting with normal electrocardiography and began cycling for 5 min at 60 to 70 revolutions per minute using a light external resistance, as determined according to body weight. The external resistance was gradually increased by 25 watts per minute until voluntary exhaustion. The W_Wax_ has been shown to have a strong correlation (r = 0.88) with maximal oxygen uptake (i.e., the gold standard metric of cardiorespiratory fitness in young people.^24^ We classified recorded values of <100 W_max_ as extremely low, and subsequently excluded these conscripts as in previous studies. ^25,26^

### Ascertainment of type 2 diabetes

The outcome was T2D defined as a composite of either a diagnosis or dispensation of antidiabetic medication until 31 December 2023. The combination of diagnoses and dispensation of medications in the ascertainment of T2D has previously been used in Swedish registry studies.^27^ Diagnoses were ascertained using the National Patient Register and the International Classification of Diseases, 10^th^ revision (ICD-10), code E11. Inpatient diagnosis of T2D in the National Patient Register has a positive predictive value of 79% to 100%, although sensitivity is lower (23% to 84%).^20^ Dispensation of antidiabetic medication in the Prescribed Drug Register was collected using the Anatomical Therapeutic Chemical code A10.^21^

### Covariates

Data on age at conscription, year of conscription, and objectively measured body mass index (kg/m^2^) were obtained from the Swedish Military Conscription Register. We considered values ≤15 and ≥60 kg/m^2^ as extreme and subsequently excluded those with such values, as in previous studies.^25,26^ We collected socioeconomic data of both mothers and fathers of the conscripts from databases managed by Statistics Sweden.^23^ Specifically, we obtained information on the highest attained lifetime education and the annual disposable income standardised by birth years into highest-achieved quintiles between ages 40 and 50. This approach was chosen in an attempt to capture working life income and is henceforth referred to as income categories. When values for both parents were available, we retained the highest value.

### Statistical analysis

Participants were followed from conscription date until the date of T2D diagnosis or dispensation of antidiabetic medication, death, emigration, or end of follow-up (31 December 2023), whichever came first. Flexible parametric survival models were used to calculate hazard ratios (HRs) for T2D during follow-up by levels of fitness, with baseline knots placed at the 5^th^, 27.5^th^, 50^th^, 72.5^th^ and 95^th^ percentile of the uncensored log survival times, and using age as the underlying timescale.^28,29^ We modelled cardiorespiratory fitness both as deciles and using restricted cubic splines with knots placed at the 5^th^, 35^th^, 65^th^, and 95^th^ percentile.^29^ To enhance the clinical interpretation and utility of the results, we also computed the standardized cumulative incidence of T2D at 65 years of age (1-Survival) and illustrated it graphically over the follow-up period.^28^ Moreover, we calculated the preventable fraction of T2D at 65 years of age associated with different hypothetical public health interventions, including a “minor” (shifting everyone in the first decile of fitness to the second decile), “moderate” (shifting everyone in the bottom four deciles to the fifth decile), and “extreme” (shifting everyone to the tenth decile) intervention.^30^ All models were adjusted for age at conscription (continuous), year of conscription (1972, 1973-1977, 1978-1982, 1983-1987, 1988-1992, and 1993-1995), body mass index (continuous and quadratic term), parental education (compulsory school ≤9 years, secondary education, post-secondary education <3 years, post-secondary education ≥3 years), and parental income (5 categories). Additionally, to explore whether body mass index acted as an effect modifier, the association was also examined by strata of overweight status (above or below 25.0kg/m^2^).

For the full-sibling analysis, the flexible parametric model was extended to a marginalized between-within model with robust standard errors,^16,31,32^ which enables control for all unobserved confounders shared between siblings, including behavioral, environmental, and ≈50% of genetic factors. The between-within model includes an individual term for the exposure/covariate and a term for its family average, thereby isolating the individual-level variation (within effect) from the family-level variation (between effect).^16,31,32^

#### Sensitivity analyses

We repeated the main analysis after additional adjustment for muscular strength,^13^ and after adjustment for height.^33^ We also repeated the analysis after scaling W_max_ to body weight (W_max_/kg), and after estimating VO_2max_ from W_max_ and body weight using a validated equation.^34^ To explore whether changes in estimates from cohort to full-sibling analysis were more likely due to selection bias in the sibling cohort rather than control for unobserved confounders,^32^ the standard cohort analysis was performed in the sibling cohort (that is, without controlling for shared factors). Because medications used in the treatment of T2D can also be used for other conditions (e.g., polycystic ovary syndrome, obesity, heart failure, type 1 diabetes), and information about the indication is not available in the Prescribed Drug Register,^21^ we also repeated the main analysis but only considered patient registry recorded T2D diagnosis as the outcome. Finally, because ICD-10 was not implemented officially until 1997, leaving a possibility for conscripts from the earlier cohorts to have been followed for many years before having the possibility to receive a T2D diagnosis, we repeated the analyses restricted to those conscripted in the year 1985 or later. All analyses were performed using Stata MP version 16.1.

## Results

### Baseline characteristics

Baseline characteristics in the total cohort and in the full-sibling cohort were similar (Table 1). The mean (standard deviation) age at conscription was 18.3 (0.7) years. Most (81.3%) had a body mass index in the normal-weight range. About a quarter (26.6%) had parents with a high (post-secondary) level of education and about a third (34.1%) had parents with a high (top category) level of annual income. Compared to those with the lowest fitness levels, those with higher fitness levels were on average born later, had a higher body mass index, and had parents with a higher level of education and income (Supplemental Table 1).

**Table 1.** Baseline characteristics.

|  | **Total cohort**<br>**(N=1 124 049)** | **Full-sibling cohort**<br>**(N=477 453)** |
| --- | --- | --- |
| **Birth year, median (IQR)** | 1966 (1960 to 1971) | 1965 (1961 to 1970) |
| **Age at conscription, mean (SD)** | 18.3 (0.7) | 18.3 (0.7) |
| **Body mass index, kg/m<sup>2</sup>** |  |  |
| Mean (SD) | 21.8 (2.9) | 21.7 (2.8) |
| **Body mass index categories, n (%)** |  |  |
| Underweight (<18.5 kg/m<sup>2</sup>) | 88 966 (7.9) | 37 884 (7.9) |
| Normal weight (18.5-24.9 kg/m<sup>2</sup>) | 913 586 (81.3) | 390 811 (81.9) |
| Overweight (25.0-29.9 kg/m<sup>2</sup>) | 102 267 (9.1) | 41 248 (8.6) |
| Obesity (≥30.0 kg/m<sup>2</sup>) | 19 230 (1.7) | 7510 (1.6) |
| **Cardiorespiratory fitness, Watt max** |  |  |
| Mean (SD) | 277 (52) | 277 (52) |
| **Parental level of education, n (%)** |  |  |
| Compulsory school ≤9 years | 319 242 (28.4) | 139 246 (29.1) |
| Secondary education | 505 510 (45.0) | 210 246 (44.0) |
| Post-secondary education <3 years | 123 757 (11.0) | 50 210 (10.5) |
| Post-secondary education ≥3 years | 175 540 (15.6) | 77 751 (16.3) |
| **Parental highest income, n (%)** |  |  |
| Category 1 (low income) | 55 055 (4.9) | 18 918 (4.0) |
| Category 2 | 108 836 (9.7) | 41 820 (8.8) |
| Category 3 | 237 953 (21.2) | 101 150 (21.2) |
| Category 4 | 338 444 (30.1) | 145 735 (30.5) |
| Category 5 (high income) | 383 761 (34.1) | 169 830 (35.6) |

IQR = interquartile range. SD = standard deviation.

### Type 2 diabetes during follow-up

During follow-up, 115 958 (10.3%) in the total cohort and 48 089 (10.1%) of the full siblings were diagnosed with T2D or dispensed antidiabetic medication. In the total cohort, the median (IQR) age at first event was 53.4 (47.6 to 59.3) years (minimum 21.0; maximum 73.0). In the full-sibling cohort, the median (IQR) age at first event was 53.5 (47.8 to 59.1) years (minimum 21.0; maximum 72.5). Most cases (93.4%) occurred after the age of 40, and about two thirds (65.0%) occurred after the age of 50. The numbers censored are shown in Supplemental Table 2.

### Cardiorespiratory fitness and type 2 diabetes

In cohort analysis after accounting for covariates, higher levels of cardiorespiratory fitness were associated with a progressively lower risk of T2D, starting already from the second decile of fitness (Figure 1, Figure 2, Table 2). Compared to the first decile, the HR in the second decile was 0.83 (95% confidence interval [CI] 0.81 to 0.85), with a difference in the standardized cumulative incidence at age 65 of 4.3 (3.8 to 4.8) percentage points. From thereon, the risk gradually declined, where the HR in the tenth decile was 0.38 (0.36 to 0.39), with an incidence difference of 17.8 (17.3 to 18.3) percentage points. A minor intervention shifting everyone in the first decile of fitness to the second decile was estimated to prevent 7.2% (6.4 to 8.0) of T2D cases at age 65, whereas an extreme intervention shifting everyone to the tenth decile was estimated to prevent 35.6% (34.1 to 37.0) of cases (Supplemental Table 3). The association appeared to be similar in those with overweight compared to those without overweight (Figure 3, Supplemental Table 4).

**Figure 1.**
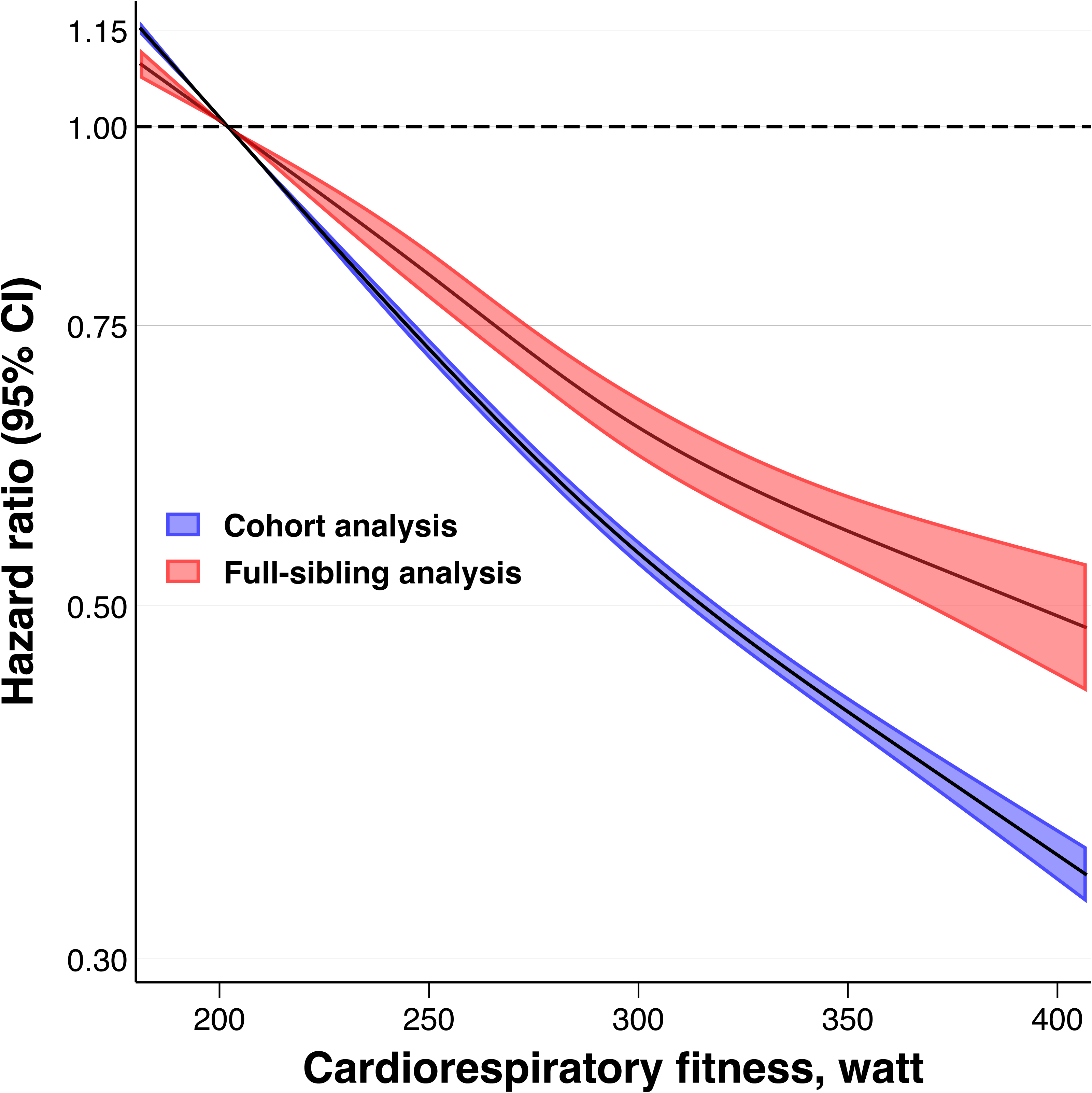
Hazard ratio for type 2 diabetes across restricted cubic splines of cardiorespiratory fitness in cohort and full-sibling analysis. Estimates were obtained using flexible parametric survival models, extended to a marginalized between-within model in the full-sibling cohort, with knots placed at the 5^th^, 35^th^, 65^th^, and 95^th^ percentile, and using age as the underlying time scale. The referent was set to the median value of the bottom decile (202 W_max_). The models were adjusted for age at conscription, year of conscription, body mass index, parental education, and parental income. For graphical purposes, the x-axis was limited to span from the first to the 99th percentile of the exposure distribution.

**Figure 2.**
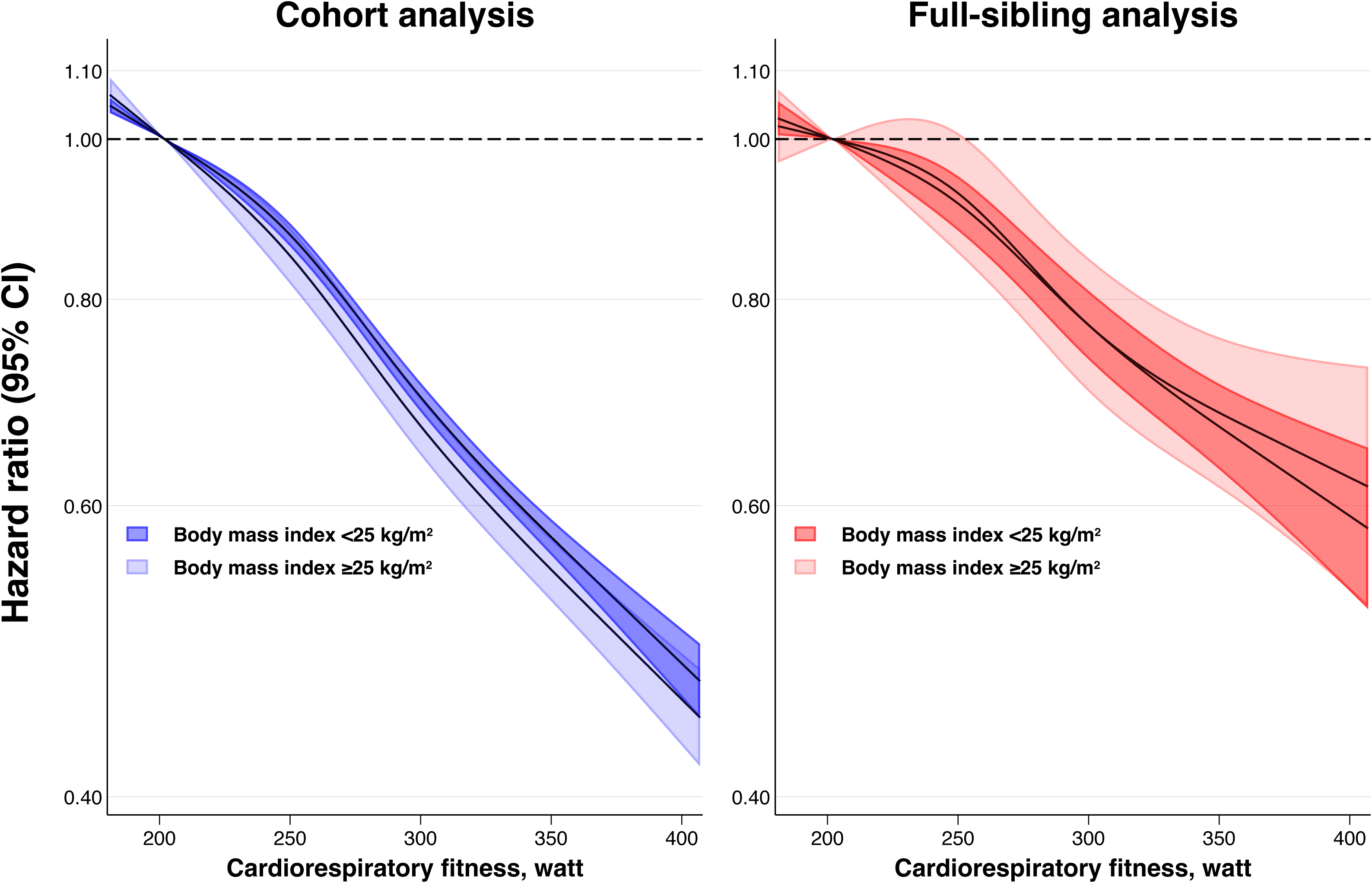
Standardized cumulative incidence for type 2 diabetes across the entire follow-up (top) and differences at age 65 (bottom) by deciles of cardiorespiratory fitness in cohort and full-sibling analysis. Estimates obtained using flexible parametric survival models, extended to a marginalized between-within model in the full-sibling cohort, with baseline knots placed at the 5^th^, 27.5^th^, 50^th^, 72.5^th^, and 95^th^ percentile of the uncensored log survival times, and using age as the underlying time scale. The bottom decile (lowest fitness) was the referent. All models were adjusted for age at conscription, year of conscription, body mass index, parental education, and parental income. Inferential measures for the incidence curves were omitted for clarity as they are reflected in the bottom panel of this figure as well as in Figure 1 and Table 2.

**Figure 3.**
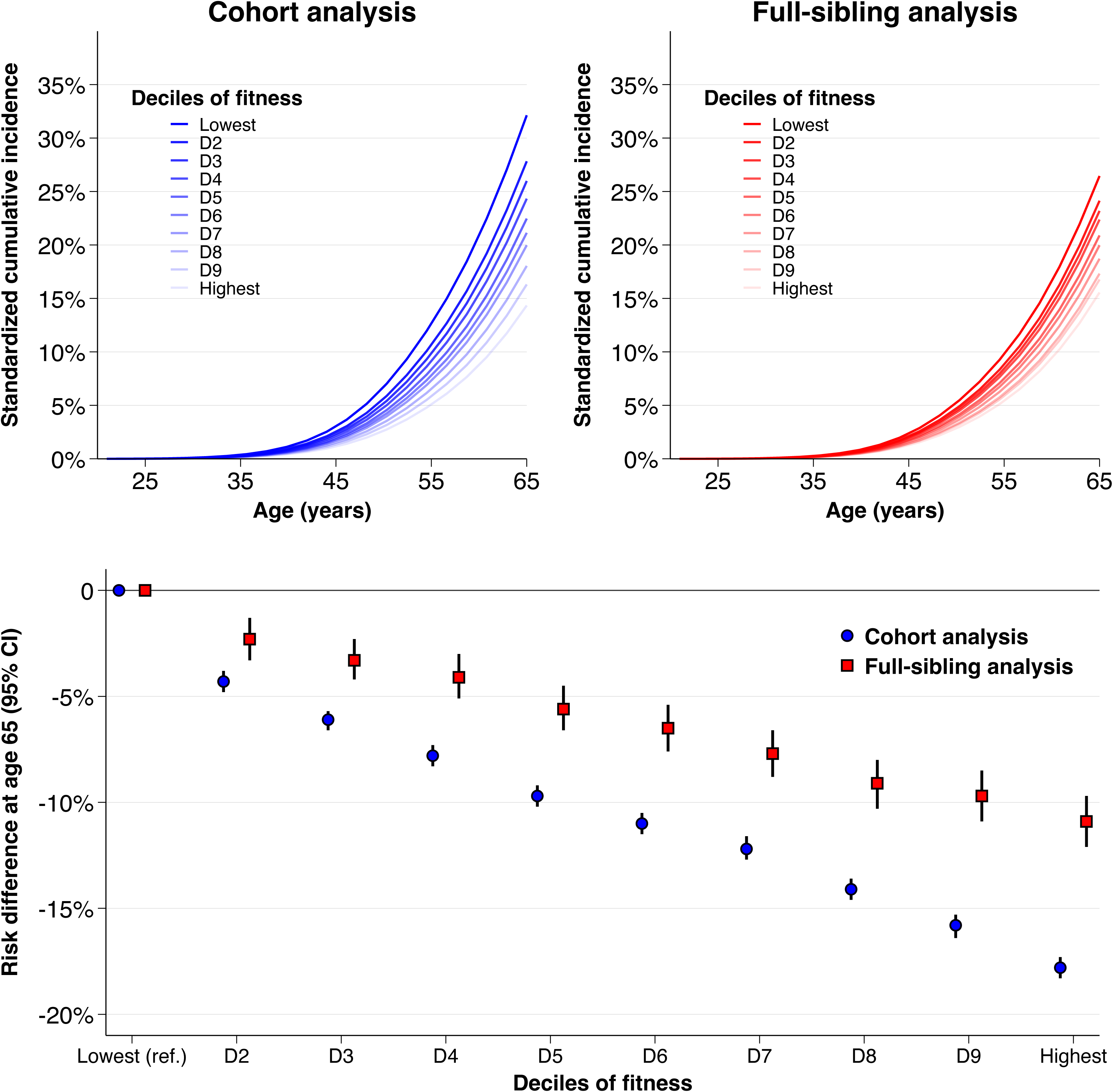
Hazard ratio for type 2 diabetes across restricted cubic splines of cardiorespiratory fitness separately in those without overweight and those with overweight, in cohort and full-sibling analysis. Estimates were obtained using flexible parametric survival models, extended to a marginalized between-within model in the full-sibling cohort, with knots placed at the 5^th^, 35^th^, 65^th^, and 95^th^ percentile, and using age as the underlying time scale. The referent was set to the median value of the bottom decile (202 W_max_). The models were adjusted for age at conscription, year of conscription, parental education, and parental income. For graphical purposes, the x-axis was limited to span from the first to the 99th percentile of the exposure distribution.

**Table 2.**
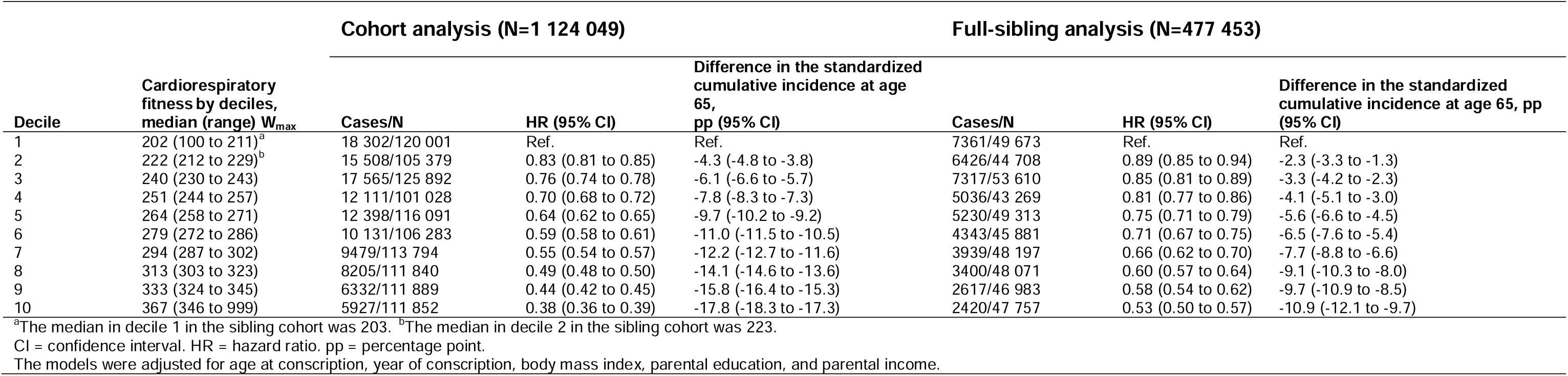
Hazard ratios for type 2 diabetes and differences in the standardized cumulative incidence at 65 years of age by deciles of cardiorespiratory fitness in cohort and full-sibling analysis.

In full-sibling analysis, the association replicated, although with attenuation in the magnitude of association (Figure 1, Figure 2, Table 2). Compared to the first decile, the HR in the second decile was 0.89 (0.85 to 0.94), with an incidence difference of 2.3 (1.3 to 3.3) percentage points. In the top decile, the HR was 0.53 (0.50 to 0.57) with an incidence difference of 10.9 (9.7 to 12.1) percentage points. A minor intervention was estimated to prevent 4.6% (2.6 to 6.5) of T2D cases at age 65, whereas an extreme intervention was estimated to prevent 24.3% (20.5 to 28.0) of cases (Supplemental Table 3).

### Sensitivity analyses

The results were not materially influenced by adjustment for muscular strength or height, or when rescaling the exposure (Supplemental Tables 5-8). Applying the standard analysis in the full-sibling cohort produced similar estimates as in the cohort analysis (Supplemental Table 9). Restring the analysis to those who conscribed 1985 or later, or using an outcome definition that only included T2D diagnoses, yielded overall similar results as in the main analysis (Supplemental Table 10-11, Supplemental Figure 2).

## Discussion

In this nationwide cohort study spanning six decades, higher levels of cardiorespiratory fitness in late adolescence were associated with a progressively lower risk of T2D in late adulthood, with clinically relevant associations starting already from low levels of fitness, and similarly in those with overweight as in those without. The association replicated after adjusting for unobserved behavioral, environmental, and genetic confounders shared between full siblings, but the magnitude of association attenuated.

Comparing the second to the first decile of fitness, for which the difference in fitness levels (W_max_) was less than 10%, there was a 4.3 percentage point difference in the standardized cumulative incidence of T2D at age 65, which translated into a hypothetically preventable fraction of about 7%. Interestingly, there was little evidence that the association leveled off at higher fitness levels. Comparing the tenth to the first decile showed a 17.8 percentage point incidence difference, and shifting everyone to the tenth decile was estimated to prevent about one third of cases.

While previous studies have typically measured cardiorespiratory fitness in middle-age and with limited follow-up,^13^ our study provides new evidence on the association across the life course, suggesting that cardiorespiratory fitness already in late adolescence is a robust non-invasive biomarker of long-term T2D risk, which may render support to large-scale surveillance initiatives.^35^ Our findings build upon those from a previous study based on the same population which also estimated strong associations between adolescent cardiorespiratory fitness and future T2D.^14^ An important strength of our study is that we examined the currently unknown role of familial confounding, while also following a larger proportion of our study population into the ages where T2D is typically diagnosed.^36^

A critical finding of this study was therefore that the association replicated after adjusting for unobserved familial confounders shared between full siblings, but with an attenuation in magnitude. Similar comparable studies are few, and the findings are inconsistent. For example, in one study, twin members who were physically active had a lower risk of T2D compared to their inactive co-twin (HR 0.54; 0.37 to 0.78).^37^ The results tended to be similar in a small sample of monozygotic twins, although with a greater degree of uncertainty due to limited statistical power.^37^ There have also been two studies using a Mendelian randomization framework, which in theory can be more robust to confounding than conventional observational analysis. One of these did not link genetically predicted cardiorespiratory fitness with T2D,^38^ whereas the other one found that higher fitness was associated with lower T2D in both conventional observational analysis and Mendelian randomization.^39^ Until now, it has remained challenging to interpret these findings for public health, considering the lack of measures that can more easily be contextualized, such as standardized incidences. In this respect, our study adds important evidence, as the difference in magnitude of association between cohort and full-sibling analysis seemed clinically relevant. For example, the incidence differences between deciles of fitness were about 40% smaller in sibling-comparison analyses as compared to cohort analysis, and the preventable share of cases associated with hypothetical interventions of different intensities was reduced by about one-third. We might assume that this to some degree reflects control for shared familial confounding, which could be supported by genetically informed studies, such as those using linkage disequilibrium score regression and showing genetic correlations between cardiorespiratory fitness and T2D risk factors,^40^ and between physical activity and T2D,^41^ which may suggest genetic and environmental confounding. Together, our findings therefore suggest that caution is warranted when relying on conventional observational studies for answering causal questions about the role of cardiorespiratory fitness in T2D, as doing so may yield inflated estimates.

The implications of this study can be further contextualized by considering evidence from randomized trials, in which the efficacy of combined lifestyle interventions against T2D incidence has been demonstrated in individuals with elevated T2D risk.^42^ However, the potential benefits of increased physical activity and/or fitness alone, even in the general population, remain unclear. Our findings indicate that fitness alone may play an important role in T2D prevention in the general population, and to a similar extent in those with vs. without overweight. The latter is worth highlighting given the rising prevalence of overweight and obesity, including in young people, which represents a key contributor to the T2D burden.^1,3,43^

### Limitations and strengths

The findings of this study should be considered in light of some limitations. First, because only men were included, generalizability to women is uncertain. There has been some evidence that the association between cardiorespiratory fitness and T2D may be slightly stronger in women than in men.^13^ Second, although we controlled for observed confounders such as body mass index and socioeconomic factors, as well as unobserved familial confounders through sibling-analysis, it is possible that residual and unobserved confounding still remains, such as from non-shared genes and behaviors. Future studies employing other types of causal analysis would therefore be valuable. Fourth, the sibling-comparison design also hinges on assumptions, where estimates that attenuate towards the null may be caused by amplified bias from non-shared confounding, measurement error, or control for shared mediators.^32,44^ Fifth, we identified T2D diagnoses after the ICD-10 was implemented (1997-), meaning that there was likely some degree of overestimation of age of first diagnosis in our study, as patients treated early on will remain uncaptured until the data became available to us. Yet, the results were similar in analyses restricted to participants from the later conscription cohorts, and such left-truncation of the T2D data is unlikely to have an influence on the different estimates in cohort vs. sibling analysis. Sixth, as the National Patient Register does not cover primary care, there is a risk of differential misclassification bias if those who were diagnosed in primary care (and not included in our study) had a better health status (such as higher fitness), and/or a less severe form of T2D, thereby potentially overestimating the association. We attempted to capture some diagnoses set in primary care by using nationwide data on dispensed antidiabetic medications, although the Prescribed Drug Register was not implemented until 2005. As shown, the associations were similar for the composite outcome of T2D diagnosis or antidiabetic medication and only T2D diagnosis.

There are also several strengths of this study, including the large sample of young men with standardized measures of cardiorespiratory fitness and prospective follow-up across several decades using nationwide registers with virtually zero attrition. Moreover, we had substantial statistical power which enabled us to perform a comprehensive set of analyses, including sibling-comparisons in a large sample of full siblings.

### Conclusions

This study suggests that higher levels of cardiorespiratory fitness in late adolescence are associated with a progressively lower risk of T2D in late adulthood, with clinically relevant associations starting already at low levels of fitness, and to a similar magnitude in those with normal weight and those with overweight. The association appears strong after adjustment for unobserved familial confounders shared between full siblings, but the magnitude of association attenuates. These findings highlight early-life cardiorespiratory fitness as a robust marker of long-term T2D risk, but that conventional observational analysis may yield biased estimates.

## Data sharing

The data in this study are not available to the public and will not be shared according to regulations under Swedish law. Researchers interested in obtaining the data may seek ethical approvals and inquire through Statistics Sweden. For further advice see: https://www.scb.se/en/services/guidance-for-researchers-and-universities/.

## Contributors

MB and PN had full access to all the data in the study and take responsibility for the integrity of the data and the accuracy of the data analysis. MB and PN conceived the study. All authors contributed to the design of the study. PN obtained the ethical approval and data. PN and VHA performed data managing. MB performed the statistical analyses. PN verified the underlying data. MB drafted the manuscript. All authors interpreted the results, critically revised the article for important intellectual content, read and approved the final version, and had final responsibility for the decision to submit for publication.

