## Supplemental material for "Cardiorespiratory Fitness in Adolescence and Risk of Type 2 Diabetes in Late Adulthood: A Nationwide Sibling-Controlled Cohort Study"

Marcel Ballin, PhD[1,2] (ORCID 0000-0002-9638-7208), Viktor H. Ahlqvist, PhD[1,3,4] (ORCID 0000-0003-1383-3194), Daniel Berglind, PhD[2,5,6] (ORCID 0000-0003-0616-7779), Mattias Brunström, MD, PhD[7] (ORCID 0000-0002-7054-0905), Angel Herraiz-Adillo, PhD[8] (ORCID 0000-0002-2691-0315), Pontus Henriksson, PhD[8](ORCID 0000-0003-2482-7048), Martin Neovius, PhD[9] (ORCID (0000-0003-2300-3055), Francisco B. Ortega, PhD[10,11] (ORCID 0000-0003-2001-1121), Anna Nordström, MD, PhD[12,13] (ORCID 0000-0003-3534-456X), Peter Nordström, MD, PhD[1] (ORCID (0000-0003-2924-508X)

[1]Department of Public Health and Caring Sciences, Clinical Geriatrics, Uppsala University, Uppsala, Sweden. [2]Centre for Epidemiology and Community Medicine, Region Stockholm, Stockholm, Sweden. [3]Department of Biomedicine, Aarhus University, Aarhus, Denmark. [4]Institute of Environmental Medicine, Karolinska Institutet, Stockholm, Sweden. [5]Department of Global Public Health, Karolinska Institutet, Stockholm, Sweden. [6]Center for Wellbeing, Welfare and Happiness, Stockholm School of Economics, Stockholm, Sweden. [7]Department of Public Health and Clinical Medicine, Umeå University, Umeå, Sweden. [8]Department of Health, Medicine and Caring Sciences, Linköping University, Linköping, Sweden. [9]Department of Medicine, Clinical Epidemiology Division, Karolinska Institutet, Stockholm, Sweden. [10]Department of Physical Education and Sports, Faculty of Sport Sciences, Sport and Health University Research Institute (iMUDS), University of Granada; CIBEROBN, ISCIII, Granada, Andalucía, Spain. [11]Faculty of Sport and Health Sciences, University of Jyväskylä, Jyväskylä, Finland. [12]Department of Medical Sciences, Rehabilitation Medicine, Uppsala University, Uppsala, Sweden. [13]School of Sport Sciences, UiT, The Arctic University of Norway, Tromsø, Norway

#### Correspondence:

Marcel Ballin. Department of Public Health and Caring Sciences, Clinical Geriatrics, Uppsala University, Husargatan 3 BMC, SE 75122, Uppsala, Sweden.

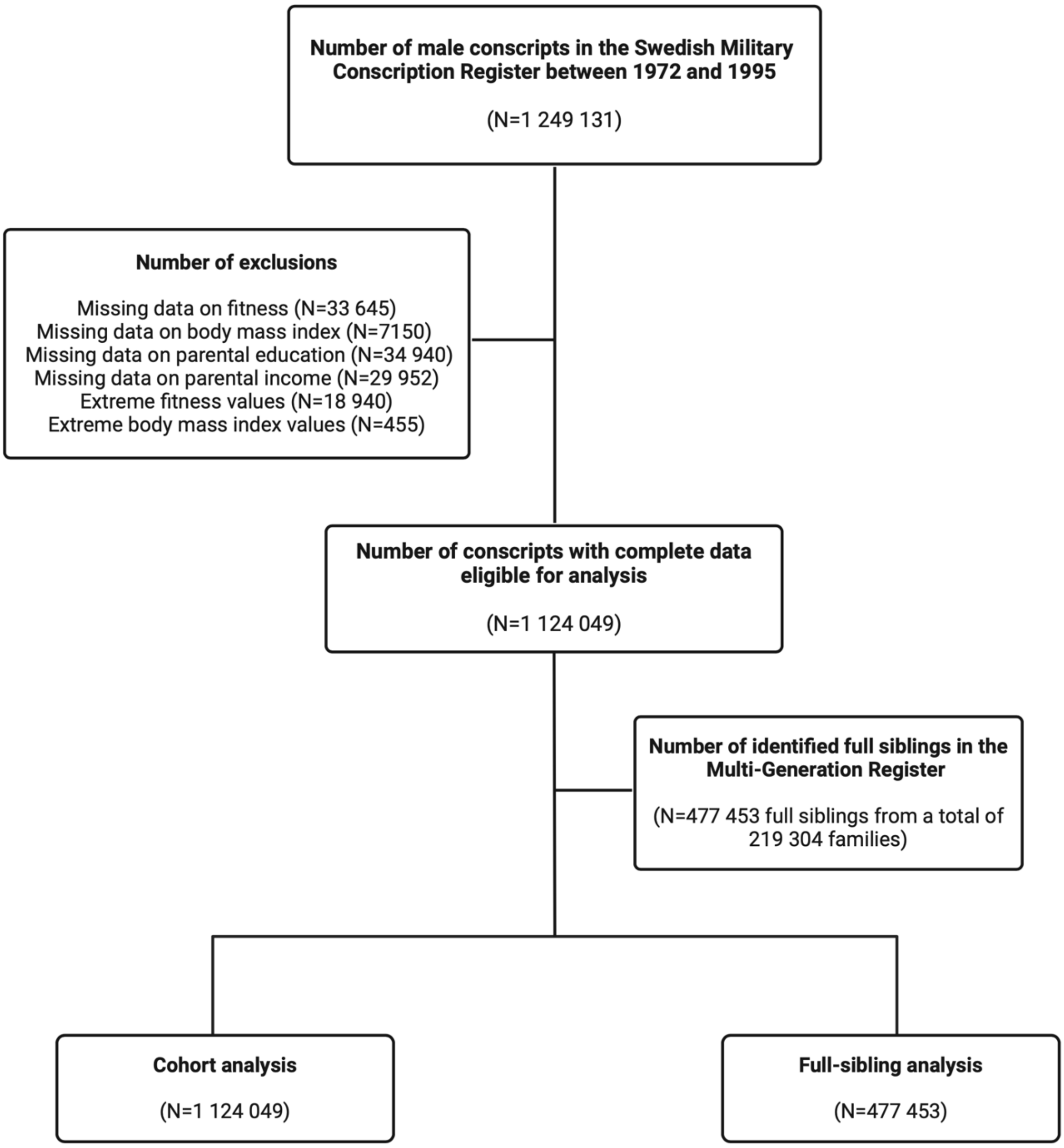


**Supplemental Figure 1. Participant flow chart.** Created using BioRender.com

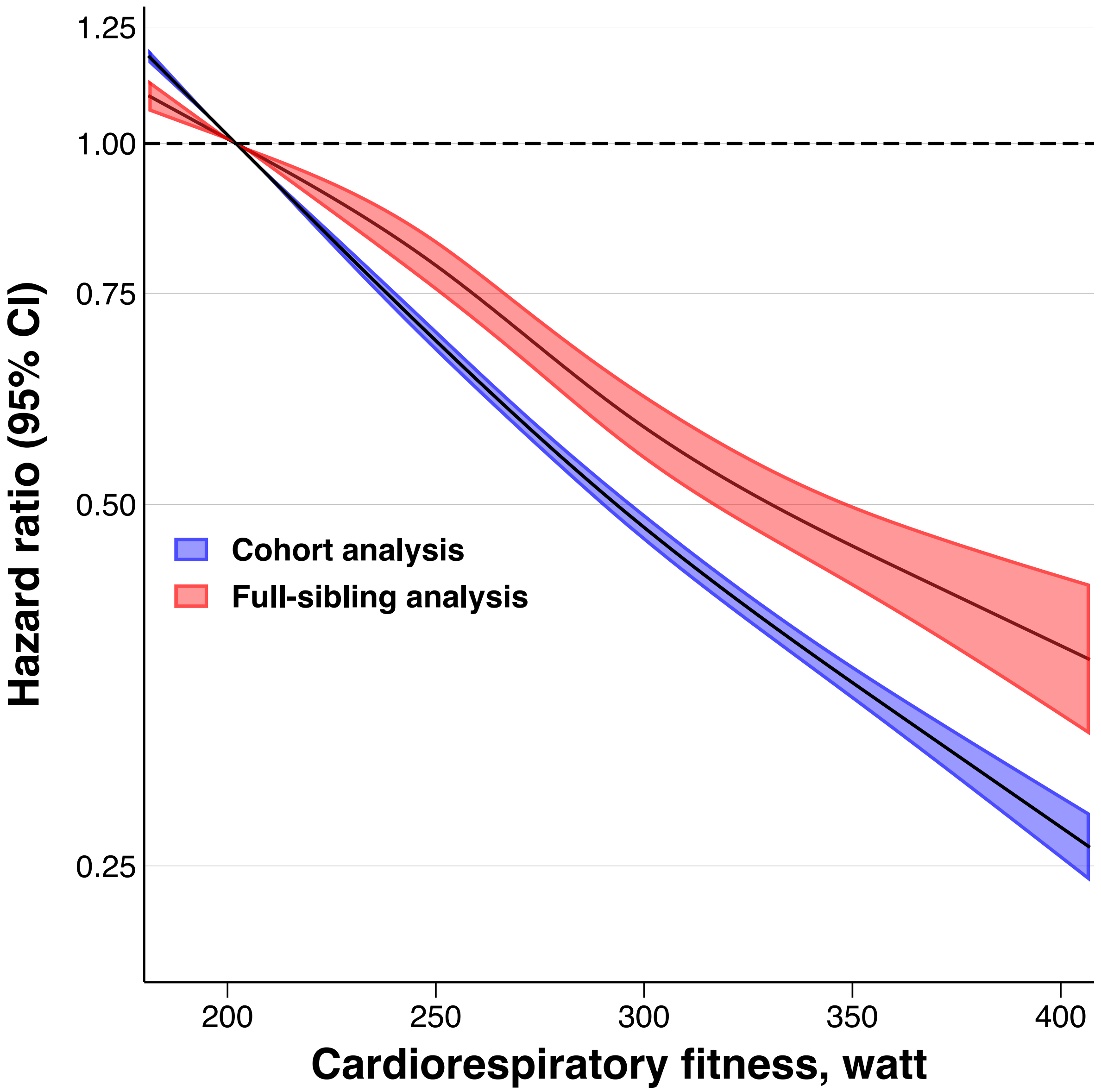


**Supplemental Figure 2. Hazard ratios for type 2 diabetes diagnosis only (thus not considering dispensation of antidiabetic medication as in the main analysis) across restricted cubic splines of cardiorespiratory fitness in cohort and full-sibling analysis.** Estimates were obtained using flexible parametric survival models, extended to a marginalized between-within model in the sibling analysis, with knots placed at the 5$^{th}$, 35$^{th}$, 65$^{th}$, and 95$^{th}$ percentile, and using age as the underlying time scale. The referent was set to the median value of the bottom decile (202 $W_{max}$). The models were adjusted for age at conscription, year of conscription, body mass index, parental education, and parental income. For graphical purposes, the x-axis was limited to span from the first to the 99th percentile of the exposure distribution.

**Supplemental Table 1. Baseline characteristics by deciles of cardiorespiratory fitness in the total cohort and in the full-sibling cohort.**

**Total cohort**

| Variables | Decile 1 (N=120 001) | Decile 2 (N=105 379) | Decile 3 (N=125 892) | Decile 4 (N=101 028) | Decile 5 (N=116 091) | Decile 6 N=106 283) | Decile 7 (N=113 794) | Decile 8 (N=111 840) | Decile 9 (N=111 889) | Decile 10 (N=111 852) |
|---|---|---|---|---|---|---|---|---|---|---|
| **Birth year, median (IQR)** | 1959 (1950-1977) | 1960 (1950-1977) | 1962 (1950-1977) | 1964 (1950-1977) | 1966 (1950-1977) | 1966 (1950-1977) | 1969 (1951-1977) | 1969 (1951-1977) | 1971 (1951-1977) | 1971 (1952-1977) |
| **Age at conscription, mean (SD)** | 18.4 (0.9) | 18.4 (0.8) | 18.3 (0.8) | 18.3 (0.7) | 18.3 (0.7) | 18.3 (0.6) | 18.3 (0.6) | 18.3 (0.6) | 18.3 (0.5) | 18.3 (0.5) |
| **Body mass index, $kg/m^2$, mean (SD)** | 20.2 (2.8) | 21.0 (2.7) | 21.4 (2.8) | 21.6 (2.9) | 21.8 (2.8) | 21.9 (2.7) | 22.1 (2.8) | 22.4 (2.7) | 22.3 (2.5) | 23.1 (2.6) |
| **Body mass index categories, n (%)** | | | | | | | | | | |
| Underweight (<18.5 $kg/m^2$) | 30 428 (25.4) | 14 233 (13.5) | 12 332 (9.8) | 8486 (8.4) | 7540 (6.5) | 5690 (5.4) | 4560 (4.0) | 2782 (2.5) | 2300 (2.1) | 615 (0.6) |
| Normal weight (18.5-24.9 $kg/m^2$) | 82 973 (69.1) | 83 468 (79.2) | 101 670 (80.8) | 82 565 (81.7) | 96 076 (82.8) | 88 964 (83.7) | 95 128 (83.6) | 94 443 (84.4) | 96 551 (86.3) | 91 748 (82.0) |
| Overweight (25.0-29.9 $kg/m^2$) | 5149 (4.3) | 6319 (6.0) | 9832 (7.8) | 8182 (8.1) | 10 399 (9.0) | 9848 (9.3) | 11 708 (10.3) | 12 629 (11.3) | 11 312 (10.1) | 16 889 (15.1) |
| Obesity (≥30.0 $kg/m^2$) | 1451 (1.2) | 1359 (1.3) | 2058 (1.6) | 1795 (1.8) | 2076 (1.8) | 1781 (1.7) | 2398 (2.1) | 1986 (1.8) | 1728 (1.5) | 2600 (2.3) |
| **Parental level of education, n (%)** | | | | | | | | | | |
| Compulsory school ≤9 years | 51 207 (42.7) | 42 796 (40.6) | 46 625 (37.0) | 32 667 (32.3) | 34 298 (29.5) | 28 527 (26.8) | 25 999 (22.9) | 22 642 (20.2) | 18 587 (16.6) | 15 894 (14.2) |
| Secondary education | 50 620 (42.2) | 44 854 (42.6) | 54 941 (43.6) | 45 737 (45.3) | 53 143 (45.8) | 49 235 (46.3) | 53 597 (47.1) | 51 869 (46.4) | 51 750 (46.3) | 49 764 (44.5) |
| Post-secondary education <3 years | 8336 (7.0) | 7932 (7.5) | 10 647 (8.5) | 9831 (9.7) | 12 291 (10.6) | 11 836 (11.1) | 14 190 (12.5) | 14 934 (13.4) | 16 394 (14.7) | 17 366 (15.5) |
| Post-secondary education ≥3 years | 9838 (8.2) | 9797 (9.3) | 13 679 (10.9) | 12 793 (12.7) | 16 359 (14.1) | 16 685 (15.7) | 20 008 (17.6) | 22 395 (20.0) | 25 158 (22.5) | 28 828 (25.8) |
| **Parental highest income, n (%)** | | | | | | | | | | |
| Category 1 (low income) | 8341 (7.0) | 6828 (6.5) | 7434 (5.9) | 5615 (5.6) | 5911 (5.1) | 5058 (4.8) | 5009 (4.4) | 4127 (3.7) | 3635 (3.3) | 3097 (2.8) |
| Category 2 | 14 583 (12.2) | 11 914 (11.3) | 13 719 (10.9) | 10 537 (10.4) | 11 770 (10.1) | 10 081 (9.5) | 10 425 (9.2) | 9344 (8.4) | 8928 (8.0) | 7534 (6.7) |
| Category 3 | 30 064 (25.1) | 25 264 (24.0) | 28 880 (22.9) | 22 528 (22.3) | 25 028 (21.6) | 22 293 (21.0) | 23 274 (20.5) | 21 200 (19.0) | 20 635 (18.4) | 18 787 (16.8) |
| Category 4 | 36 612 (30.5) | 31 776 (30.1) | 38 634 (30.7) | 30 654 (30.3) | 35 124 (30.3) | 32 279 (30.4) | 34 044 (29.9) | 33 159 (29.7) | 33 252 (29.7) | 32 921 (29.4) |
| Category 5 (high income) | 30 401 (25.3) | 29 597 (28.1) | 37 225 (29.6) | 31 694 (31.4) | 38 258 (33.0) | 36 572 (34.4) | 41 053 (36.1) | 44 010 (39.4) | 45 438 (40.6) | 49 513 (44.3) |

**Full-sibling cohort**

| | Decile 1 (N=49 673) | Decile 2 (N=44 708) | Decile 3 (N=53 610) | Decile 4 (N=43 260) | Decile 5 (N=49 313) | Decile 6 (N=45 881) | Decile 7 (N=48 197) | Decile 8 (N=48 071) | Decile 9 (N=46 983) | Decile 10 (N=47 757) |
|---|---|---|---|---|---|---|---|---|---|---|
| **Birth year, median (IQR)** | 1961 (1950-1977) | 1961 (1950-1977) | 1962 (1950-1977) | 1964 (1950-1977) | 1965 (1951-1977) | 1966 (1950-1977) | 1968 (1952-1977) | 1968 (1952-1977) | 1970 (1952-1977) | 1970 (1953-1977) |
| **Age at conscription, mean (SD)** | 18.4 (0.9) | 18.3 (0.8) | 18.3 (0.8) | 18.3 (0.7) | 18.3 (0.7) | 18.3 (0.6) | 18.3 (0.6) | 18.2 (0.5) | 18.3 (0.5) | 18.3 (0.5) |
| **Body mass index, $kg/m^2$, mean (SD)** | 20.2 (2.7) | 20.9 (2.7) | 21.4 (2.8) | 21.5 (2.8) | 21.7 (2.7) | 21.8 (2.7) | 22.1 (2.8) | 22.3 (2.6) | 22.2 (2.5) | 23.0 (2.6) |
| **Body mass index categories, n (%)** | | | | | | | | | | |
| Underweight (<18.5 $kg/m^2$) | 12 743 (25.7) | 6189 (13.8) | 5242 (9.8) | 3627 (8.4) | 3179 (6.5) | 2463 (5.4) | 1929 (4.0) | 1213 (2.5) | 1015 (2.2) | 284 (0.6) |
| Normal weight (18.5-24.9 $kg/m^2$) | 34 350 (69.2) | 35 427 (79.2) | 43 480 (81.1) | 35 554 (82.2) | 41 107 (83.4) | 38 664 (84.3) | 40 617 (84.3) | 41 035 (85.4) | 40 811 (86.9) | 39 766 (83.3) |
| Overweight (25.0-29.9 $kg/m^2$) | 2051 (4.1) | 2550 (5.7) | 4062 (7.6) | 3333 (7.7) | 4211 (8.5) | 4043 (8.8) | 4736 (9.8) | 5044 (10.5) | 4496 (10.0) | 6722 (14.1) |
| Obesity (≥30.0 $kg/m^2$) | 529 (1.1) | 542 (1.2) | 826 (1.5) | 746 (1.7) | 816 (1.7) | 711 (1.6) | 915 (1.9) | 779 (1.5) | 661 (1.4) | 985 (2.1) |
| **Parental level of education, n (%)** | | | | | | | | | | |
| Compulsory school ≤9 years | 21 446 (43.2) | 18 112 (40.5) | 19 901 (37.1) | 14 344 (33.2) | 15 096 (30.6) | 12 831 (28.0) | 11 617 (24.1) | 10 350 (21.5) | 8370 (17.8) | 7179 (15.0) |
| Secondary education | 20 714 (41.7) | 18 879 (42.2) | 23 197 (43.3) | 19 207 (44.4) | 22 077 (44.8) | 20 697 (45.1) | 22 160 (46.0) | 21 548 (44.8) | 21 195 (45.1) | 20 572 (43.1) |
| Post-secondary education <3 years | 3327 (6.7) | 3290 (7.4) | 4331 (8.1) | 4010 (9.3) | 4993 (10.1) | 4902 (10.7) | 5694 (11.8) | 6076 (12.6) | 6545 (13.9) | 7042 (14.8) |
| Post-secondary education ≥3 years | 4186 (8.4) | 4427 (9.9) | 6181 (11.5) | 5699 (13.2) | 7147 (14.5) | 7451 (16.2) | 8726 (18.1) | 10 097 (21.0) | 10 873 (23.1) | 12 964 (27.2) |
| **Parental highest income, n (%)** | | | | | | | | | | |
| Category 1 (low income) | 2759 (5.6) | 2333 (5.2) | 2469 (4.6) | 1940 (4.5) | 2008 (4.1) | 1790 (3.9) | 1767 (3.7) | 1481 (3.1) | 1264 (2.7) | 1107 (2.3) |
| Category 2 | 5579 (11.2) | 4619 (10.3) | 5358 (10.0) | 4103 (9.5) | 4494 (9.1) | 3936 (8.6) | 3995 (8.3) | 3552 (7.4) | 3303 (7.0) | 2881 (6.0) |
| Category 3 | 12 839 (25.9) | 10 890 (24.4) | 12 379 (23.1) | 9761 (22.6) | 10 757 (21.8) | 9522 (20.8) | 9838 (20.4) | 8852 (18.4) | 8444 (18.0) | 7868 (16.5) |
| Category 4 | 15 791 (31.8) | 13 803 (30.9) | 16 802 (31.3) | 13 356 (30.9) | 15 229 (30.9) | 14 200 (31.0) | 14 458 (30.0) | 14 321 (29.8) | 13 986 (29.8) | 13 789 (28.9) |
| Category 5 (high income) | 12 705 (25.6) | 13 063 (29.2) | 16 602 (31.0) | 14 100 (32.6) | 16 825 (34.1) | 16 433 (35.8) | 18 139 (37.6) | 19 865 (41.3) | 19 986 (42.5) | 22 112 (46.3) |

IQR = interquartile range. SD = standard deviation.

### Supplemental Table 2. Numbers censored due to death, emigration, and end of follow-up.

| | Cohort analysis (N=1 124 049) | Full-sibling analysis (N=477 453) |
|---|---|---|
| **During the total follow-up** | | |
| Death | 56 009 (5.0) | 22 304 (4.7) |
| Emigration | 71 656 (6.4) | 29 530 (6.2) |
| End of follow-up, 31 Dec 2023 | 880 426 (78.3) | 377 530 (79.1) |
| **Before 1997** | | |
| Death | 10 772 (1.0) | 4126 (0.9) |
| Emigration | 20 775 (1.9) | 9077 (1.9) |

Number of events and numbers censored are shown as n (%).

**Supplemental Table 3. Population-attributable fraction of type 2 diabetes at 65 years of age by cardiorespiratory fitness in cohort and full-sibling analysis, when considering a minor, (shifting those in decile 1 to decile 2), moderate (shifting those below deciles 5 to decile 5), or an extreme intervention (shifting everyone to decile 10).**

| | **Cohort analysis (N=1 124 049)** | **Full-sibling analysis (N=477 453)** |
|---|---|---|
| **Hypothetical intervention** | **PAF, % (95% CI)** | **PAF, % (95% CI)** |
| **Minor** | 7.2 (6.4 to 8.0) | 4.6 (2.6 to 6.5) |
| **Moderate** | 15.4 (14.2 to 16.5) | 10.8 (8.1 to 13.4) |
| **Extreme** | 35.6 (34.1 to 37.0) | 24.3 (20.5 to 28.0) |

CI = confidence interval. PAF = population-attributable fraction.

The models were adjusted for age at conscription, year of conscription, body mass index, parental education, and parental income.

**Supplemental Table 4. Hazard ratios for type 2 diabetes by deciles of cardiorespiratory fitness in cohort and full-sibling analysis stratified by overweight status.**

| Cardiorespiratory fitness, deciles | Cohort analysis in participants without overweight (N=1 002 552) HR (95% CI) | Cohort analysis in participants with overweight (N=121 497) HR (95% CI) | Full-sibling analysis in participants without overweight (N=428 695) HR (95% CI) | Full-sibling analysis in participants with overweight N=48 758) HR (95% CI) |
|---|---|---|---|---|
| D1 | Ref. | Ref. | Ref. | Ref. |
| D2 | 0.92 (0.90 to 0.94) | 0.98 (0.92 to 1.03) | 0.95 (0.91 to 1.01) | 1.02 (0.91 to 1.15) |
| D3 | 0.91 (0.89 to 0.93) | 0.88 (0.83 to 0.93) | 0.96 (0.90 to 1.01) | 0.97 (0.87 to 1.09) |
| D4 | 0.86 (0.84 to 0.88) | 0.85 (0.80 to 0.90) | 0.95 (0.89 to 1.00) | 0.92 (0.81 to 1.04) |
| D5 | 0.80 (0.78 to 0.82) | 0.80 (0.76 to 0.85) | 0.87 (0.82 to 0.92) | 0.92 (0.81 to 1.03) |
| D6 | 0.75 (0.73 to 0.77) | 0.74 (0.70 to 0.79) | 0.85 (0.80 to 0.90) | 0.83 (0.73 to 0.94) |
| D7 | 0.72 (0.70 to 0.75) | 0.70 (0.66 to 0.74) | 0.78 (0.73 to 0.84) | 0.81 (0.72 to 0.93) |
| D8 | 0.66 (0.64 to 0.68) | 0.62 (0.59 to 0.66) | 0.73 (0.68 to 0.79) | 0.74 (0.65 to 0.84) |
| D9 | 0.59 (0.57 to 0.61) | 0.58 (0.55 to 0.62) | 0.71 (0.66 to 0.77) | 0.74 (0.64 to 0.85) |
| D10 | 0.53 (0.50 to 0.54) | 0.50 (0.47 to 0.54) | 0.65 (0.60 to 0.71) | 0.67 (0.58 to 0.77) |

CI = confidence interval. D = decile. HR = hazard ratio.
The models were adjusted for age at conscription, year of conscription, parental education, and parental income.

**Supplemental Table 5. Hazard ratios for type 2 diabetes by deciles of cardiorespiratory fitness in cohort and full-sibling analysis, with and without adjusting for handgrip strength.**

| | **Cohort analysis without adjustment for handgrip strength (N=1 124 049)** | **Cohort analysis with adjustment for handgrip strength (N=1 095 378)** | **Full-sibling analysis without adjustment for handgrip strength (N=477 453)** | **Full-sibling analysis with adjustment for handgrip strength (N=469 718)** |
|---|---|---|---|---|
| **Cardiorespiratory fitness, deciles** | **HR (95% CI)** | **HR (95% CI)** | **HR (95% CI)** | **HR (95% CI)** |
| D1 | Ref. | Ref. | Ref. | Ref. |
| D2 | 0.83 (0.81 to 0.85) | 0.85 (0.83 to 0.87) | 0.89 (0.85 to 0.94) | 0.91 (0.86 to 0.95) |
| D3 | 0.76 (0.74 to 0.78) | 0.79 (0.77 to 0.80) | 0.85 (0.81 to 0.89) | 0.88 (0.84 to 0.92) |
| D4 | 0.70 (0.68 to 0.72) | 0.73 (0.71 to 0.75) | 0.82 (0.78 to 0.86) | 0.85 (0.80 to 0.90) |
| D5 | 0.64 (0.62 to 0.65) | 0.67 (0.65 to 0.69) | 0.76 (0.72 to 0.80) | 0.79 (0.75 to 0.84) |
| D6 | 0.59 (0.58 to 0.61) | 0.62 (0.61to 0.64) | 0.71 (0.67 to 0.76) | 0.76 (0.71 to 0.80) |
| D7 | 0.55 (0.54 to 0.57) | 0.59 (0.57 to 0.60) | 0.66 (0.62 to 0.70) | 0.71 (0.67 to 0.76) |
| D8 | 0.49 (0.48 to 0.50) | 0.53 (0.51 to 0.54) | 0.60 (0.57 to 0.64) | 0.66 (0.62 to 0.70) |
| D9 | 0.44 (0.42 to 0.45) | 0.47 (0.46 to 0.49) | 0.58 (0.54 to 0.62) | 0.64 (0.59 to 0.68) |
| D10 | 0.38 (0.36 to 0.39) | 0.42 (0.40 to 0.43) | 0.53 (0.49 to 0.57) | 0.60 (0.56 to 0.64) |

CI = confidence interval. D = decile. HR = hazard ratio.
The models were adjusted for age at conscription, year of conscription, body mass index, parental education, parental income, and handgrip strength as indicated.

**Supplemental Table 6. Hazard ratios for type 2 diabetes by deciles of cardiorespiratory fitness in cohort and full-sibling analysis, with and without adjusting for knee extension strength.**

| Cardiorespiratory fitness, deciles | Cohort analysis without adjustment for knee extension strength (N=1 124 049) | Cohort analysis with adjustment for knee extension strength (N=1 095 343) | Full-sibling analysis without adjustment for knee extension strength (N=477 453) | Full-sibling analysis with adjustment for knee extension strength (N=469 686) |
|---|---|---|---|---|
| | **HR (95% CI)** | **HR (95% CI)** | **HR (95% CI)** | **HR (95% CI)** |
| D1 | Ref. | Ref. | Ref. | Ref. |
| D2 | 0.83 (0.81 to 0.85) | 0.85 (0.83 to 0.86) | 0.89 (0.85 to 0.94) | 0.91 (0.87 to 0.95) |
| D3 | 0.76 (0.74 to 0.78) | 0.79 (0.77 to 0.80) | 0.85 (0.81 to 0.89) | 0.88 (0.84 to 0.92) |
| D4 | 0.70 (0.68 to 0.72) | 0.73 (0.71 to 0.75) | 0.82 (0.78 to 0.86) | 0.85 (0.81 to 0.90) |
| D5 | 0.64 (0.62 to 0.65) | 0.67 (0.65 to 0.69) | 0.76 (0.72 to 0.80) | 0.80 (0.76 to 0.84) |
| D6 | 0.59 (0.58 to 0.61) | 0.63 (0.61 to 0.64) | 0.71 (0.67 to 0.76) | 0.76 (0.72 to 0.81) |
| D7 | 0.55 (0.54 to 0.57) | 0.59 (0.57 to 0.61) | 0.66 (0.62 to 0.70) | 0.72 (0.68 to 0.76) |
| D8 | 0.49 (0.48 to 0.50) | 0.53 (0.51 to 0.54) | 0.60 (0.57 to 0.64) | 0.67 (0.63 to 0.71) |
| D9 | 0.44 (0.42 to 0.45) | 0.48 (0.46 to 0.49) | 0.58 (0.54 to 0.62) | 0.65 (0.61 to 0.69) |
| D10 | 0.38 (0.36 to 0.39) | 0.42 (0.40 to 0.43) | 0.53 (0.49 to 0.57) | 0.61 (0.57 to 0.67) |

CI = confidence interval. D = decile. HR = hazard ratio.
The models were adjusted for age at conscription, year of conscription, body mass index, parental education, parental income, and knee extension strength as indicated.

### Supplemental Table 7. Hazard ratios for type 2 diabetes by deciles of cardiorespiratory fitness in cohort and full-sibling analysis, with and without adjusting for body height.

| Cardiorespiratory fitness, deciles | Cohort analysis without adjustment for body height (N=1 124 049) HR (95% CI) | Cohort analysis with adjustment for body height (N=1 124 049) HR (95% CI) | Full-sibling analysis without adjustment for body height (N=477 453) HR (95% CI) | Full-sibling analysis with adjustment for body height (N=477 453) HR (95% CI) |
|---|---|---|---|---|
| D1 | Ref. | Ref. | Ref. | Ref. |
| D2 | 0.83 (0.81 to 0.85) | 0.83 (0.81 to 0.85) | 0.89 (0.85 to 0.94) | 0.89 (0.85 to 0.94) |
| D3 | 0.76 (0.74 to 0.78) | 0.76 (0.74 to 0.77) | 0.85 (0.81 to 0.89) | 0.85 (0.82 to 0.89) |
| D4 | 0.70 (0.68 to 0.72) | 0.70 (0.68 to 0.72) | 0.82 (0.78 to 0.86) | 0.82 (0.78 to 0.87) |
| D5 | 0.64 (0.62 to 0.65) | 0.63 (0.62 to 0.65) | 0.76 (0.72 to 0.80) | 0.76 (0.72 to 0.80) |
| D6 | 0.59 (0.58 to 0.61) | 0.59 (0.57 to 0.60) | 0.71 (0.67 to 0.76) | 0.72 (0.68 to 0.77) |
| D7 | 0.55 (0.54 to 0.57) | 0.55 (0.54 to 0.56) | 0.66 (0.62 to 0.70) | 0.67 (0.64 to 0.72) |
| D8 | 0.49 (0.48 to 0.50) | 0.49 (0.47 to 0.50) | 0.60 (0.57 to 0.64) | 0.62 (0.58 to 0.66) |
| D9 | 0.44 (0.42 to 0.45) | 0.43 (0.42 to 0.45) | 0.58 (0.54 to 0.62) | 0.60 (0.56 to 0.64) |
| D10 | 0.38 (0.36 to 0.39) | 0.37 (0.36 to 0.39) | 0.53 (0.49 to 0.57) | 0.55 (0.51 to 0.59) |

CI = confidence interval. D = decile. HR = hazard ratio.
The models were adjusted for age at conscription, year of conscription, body mass index, body height, parental education, parental income, and body height as indicated.

**Supplemental Table 8. Hazard ratios for type 2 diabetes by deciles of cardiorespiratory fitness expressed as $W_{max}$, $W_{max}$/kg, or estimated $VO_{2max}$ in cohort and full-sibling analysis.**

| | $W_{max}$ | | $W_{max}$/kg | | Estimated $VO_{2max}$ | |
|---|---|---|---|---|---|---|
| | **Cohort analysis (N=1 124 049)** | **Full-sibling analysis (N=477 453)** | **Cohort analysis (N=1 124 049)** | **Full-sibling analysis (N=477 453)** | **Cohort analysis (N=1 124 049)** | **Full-sibling analysis (N=477 453)** |
| **Cardiorespiratory fitness, deciles** | **HR (95% CI)** | **HR (95% CI)** | **HR (95% CI)** | **HR (95% CI)** | **HR (95% CI)** | **HR (95% CI)** |
| D1 | Ref. | Ref. | Ref. | Ref. | Ref. | Ref. |
| D2 | 0.83 (0.81 to 0.85) | 0.89 (0.85 to 0.94) | 0.85 (0.84 to 0.87) | 0.92 (0.88 to 0.97) | 0.86 (0.84 to 0.87) | 0.92 (0.88 to 0.97) |
| D3 | 0.76 (0.74 to 0.78) | 0.85 (0.81 to 0.89) | 0.80 (0.78 to 0.82) | 0.87 (0.82 to 0.91) | 0.80 (0.78 to 0.82) | 0.87 (0.82 to 0.91) |
| D4 | 0.70 (0.68 to 0.72) | 0.81 (0.77 to 0.86) | 0.74 (0.73 to 0.76) | 0.81 (0.77 to 0.86) | 0.74 (0.73 to 0.76) | 0.81 (0.77 to 0.86) |
| D5 | 0.64 (0.62 to 0.65) | 0.75 (0.71 to 0.79) | 0.68 (0.67 to 0.70) | 0.76 (0.72 to 0.80) | 0.68 (0.67 to 0.70) | 0.76 (0.72 to 0.80) |
| D6 | 0.59 (0.58 to 0.61) | 0.71 (0.67 to 0.75) | 0.63 (0.61 to 0.65) | 0.73 (0.68 to 0.77) | 0.63 (0.61 to 0.65) | 0.73 (0.68 to 0.77) |
| D7 | 0.55 (0.54 to 0.57) | 0.66 (0.62 to 0.70) | 0.56 (0.55 to 0.58) | 0.66 (0.62 to 0.70) | 0.56 (0.55 to 0.58) | 0.66 (0.62 to 0.70) |
| D8 | 0.49 (0.48 to 0.50) | 0.60 (0.57 to 0.64) | 0.52 (0.50 to 0.53) | 0.61 (0.57 to 0.65) | 0.52 (0.50 to 0.53) | 0.61 (0.57 to 0.65) |
| D9 | 0.44 (0.42 to 0.45) | 0.58 (0.54 to 0.62) | 0.43 (0.42 to 0.45) | 0.54 (0.50 to 0.58) | 0.43 (0.42 to 0.45) | 0.54 (0.50 to 0.58) |
| D10 | 0.38 (0.36 to 0.39) | 0.53 (0.50 to 0.57) | 0.33 (0.32 to 0.35) | 0.44 (0.40 to 0.48) | 0.33 (0.32 to 0.35) | 0.44 (0.40 to 0.48) |
| **Cardiorespiratory fitness, per 1 MET** | | | | | 0.88 (0.87 to 0.89) | 0.90 (0.90 to 0.91) |

CI = confidence interval. D = decile. HR = hazard ratio. MET = metabolic equivalent of task, computed from dividing estimated $VO_{2max}$ / 3.5.
The models were adjusted for age at conscription, year of conscription, body mass index, parental education, and parental income.

**Supplemental Table 9. Hazard ratios for type 2 diabetes by deciles of cardiorespiratory fitness in cohort analysis (as reported in the main article), in the sibling cohort using standard cohort analysis, and using full-sibling analysis (as reported in the main article)**

| | Cohort analysis as reported in the main article (N=1 124 049) | Standard analysis replicated in the full-sibling cohort (N=477 453) | Full-sibling analysis as reported in the main article (N=477 453) |
|---|---|---|---|
| **Cardiorespiratory fitness, deciles** | **HR (95% CI)** | **HR (95% CI)** | **HR (95% CI)** |
| D1 | Ref. | Ref. | Ref. |
| D2 | 0.83 (0.81 to 0.85) | 0.83 (0.81 to 0.86) | 0.89 (0.85 to 0.94) |
| D3 | 0.76 (0.74 to 0.78) | 0.77 (0.74 to 0.79) | 0.85 (0.81 to 0.89) |
| D4 | 0.70 (0.68 to 0.72) | 0.69 (0.70 to 0.72) | 0.81 (0.77 to 0.86) |
| D5 | 0.64 (0.62 to 0.65) | 0.64 (0.62 to 0.66) | 0.75 (0.71 to 0.79) |
| D6 | 0.59 (0.58 to 0.61) | 0.60 (0.57 to 0.62) | 0.71 (0.67 to 0.75) |
| D7 | 0.55 (0.54 to 0.57) | 0.55 (0.53 to 0.57) | 0.66 (0.62 to 0.70) |
| D8 | 0.49 (0.48 to 0.50) | 0.48 (0.46 to 0.50) | 0.60 (0.57 to 0.64) |
| D9 | 0.44 (0.42 to 0.45) | 0.43 (0.41 to 0.45) | 0.58 (0.54 to 0.62) |
| D10 | 0.38 (0.36 to 0.39) | 0.37 (0.35 to 0.39) | 0.53 (0.50 to 0.57) |

CI = confidence interval. D = decile. HR = hazard ratio. The models were adjusted for age at conscription, year of conscription, body mass index, parental education, and parental income.

### Supplemental Table 10. Hazard ratios for type 2 by deciles of cardiorespiratory fitness in cohort and full-sibling analysis, restricted to those who conscribed year 1985 or later.

| | Cohort analysis (N=529 710) | | Full-sibling analysis (N=161 663) | |
|---|---|---|---|---|
| **Cardiorespiratory fitness, deciles** | **Cases/N** | **HR (95% CI)** | **Cases/N** | **HR (95% CI)** |
| D1 | 4084/57 822 | Ref. | 1129/17 246 | Ref. |
| D2 | 3734/52 741 | 0.87 (0.83 to 0.91) | 1022/15 459 | 0.95 (0.84 to 1.07) |
| D3 | 3303/48 703 | 0.77 (0.74 to 0.81) | 909/14 309 | 0.84 (0.74 to 0.95) |
| D4 | 3982/60 694 | 0.70 (0.67 to 0.73) | 1118/18 140 | 0.75 (0.67 to 0.85) |
| D5 | 3069/48 011 | 0.64 (0.61 to 0.68) | 802/14 438 | 0.67 (0.59 to 0.77) |
| D6 | 3085/50 629 | 0.58 (0.55 to 0.61) | 860/15 507 | 0.64 (0.56 to 0.73) |
| D7 | 3061/56 475 | 0.55 (0.52 to 0.58) | 823/17 313 | 0.59 (0.52 to 0.68) |
| D8 | 2368/49 147 | 0.51 (0.48 to 0.53) | 703/15 302 | 0.63 (0.55 to 0.73) |
| D9 | 2738 53 681 | 0.46 (0.44 to 0.49) | 785/17 055 | 0.59 (0.52 to 0.68) |
| D10 | 2446/51 807 | 0.39 (0.37 to 0.41) | 691/16 894 | 0.48 (0.41 to 0.56) |

CI = confidence interval. D = decile. HR = hazard ratio. P.P = percentage point.

The models were adjusted for age at conscription, year of conscription, body mass index, parental education, and parental income.

**Supplemental Table 11. Hazard ratios for type 2 diabetes and differences in the standardized cumulative incidence at 65 years of age by deciles of cardiorespiratory fitness in cohort and full-sibling analysis, using the outcome definition that included only type 2 diabetes diagnosis.**

| | **Cohort analysis (N=1 124 049)** | | | **Full-sibling analysis (N=477 453)** | | |
|---|---|---|---|---|---|---|
| **Cardiorespiratory fitness, deciles** | **Cases/N** | **HR (95% CI)** | **Difference in the standardized cumulative incidence at age 65, pp (95% CI)** | **Cases/N** | **HR (95% CI)** | **Difference in the standardized cumulative incidence at age 65, pp (95% CI)** |
| D1 | 9775/120 001 | Ref. | Ref. | 3880/49 673 | Ref. | Ref. |
| D2 | 8261/105 379 | 0.82 (0.79 to 0.84) | -2.3 (-2.9 to -2.1) | 3357/44 708 | 0.91 (0.85 to 0.97) | -1.0 (-1.6 to -0.3) |
| D3 | 9013/125 892 | 0.72 (0.70 to 0.74) | -3.8 (-4.2 to -3.5) | 3745/53 610 | 0.83 (0.78 to 0.89) | -1.8 (-2.5 to -1.2) |
| D4 | 6035/101 028 | 0.66 (0.64 to 0.69) | -4.6 (-5.0 to -4.3) | 2469/43 260 | 0.81 (0.75 to 0.87) | -2.0 (-2.8 to -1.3) |
| D5 | 6088/116 091 | 0.60 (0.59 to 0.63) | -5.5 (-5.9 to -5.1) | 2536/49 313 | 0.72 (0.67 to 0.78) | -3.0 (-3.8 to -2.3) |
| D6 | 4614/106 283 | 0.53 (0.51 to 0.55) | -6.5 (-6.9 to -6.2) | 1980/45 881 | 0.66 (0.61 to 0.72) | -3.7 (-4.5 to -3.0) |
| D7 | 4153/113 794 | 0.49 (0.48 to 0.51) | -7.1 (-7.5 to -6.8) | 1707/48 197 | 0.60 (0.55 to 0.65) | -4.5 (-5.2 to -3.7) |
| D8 | 3404/111 840 | 0.43 (0.41 to 0.44) | -8.1 (-8.5 to -7.8) | 1391/48 071 | 0.53 (0.48 to 0.58) | -5.2 (-6.0 to -4.5) |
| D9 | 2423 111 889 | 0.37 (0.35 to 0.38) | -9.1 (-9.5 to -8.7) | 1003/46 983 | 0.49 (0.45 to 0.55) | -5.6 (-6.4 to -4.9) |
| D10 | 2193/111 852 | 0.31 (0.29 to 0.32) | -10.0 (-10.4 to -9.6) | 893/47 757 | 0.45 (0.40 to 0.50) | -6.2 (-7.0 to 5.4) |

CI = confidence interval. D = decile. HR = hazard ratio. pp = percentage point.

The models were adjusted for age at conscription, year of conscription, body mass index, parental education, and parental income.
